# High-altitude is associated with better short-term survival in critically ill COVID-19 patients admitted to the ICU

**DOI:** 10.1101/2021.01.22.21249811

**Authors:** Pablo R. Morocho Jaramillo, Katherine Simbaña-Rivera, Javier V. Velastegui Silva, Lenin Gómez-Barreno, Ana B. Ventimilla Campoverde, Juan F. Novillo Cevallos, Washington E. Almache Guanoquiza, Silvio L. Cedeño Guevara, Luis G. Imba Castro, Nelson A. Moran Puerta, Alex W. Guayta Valladares, Alex Lister, Esteban Ortiz-Prado

## Abstract

**Background:** The novel human coronavirus, SARS-CoV-2, has affected at least 218 countries worldwide. Some geographical and environmental factors are positively associated with a better or worse prognosis concerning COVID-19 disease and with lower or higher SARS-CoV-2 transmission. High altitude exposure has been associated with lower SARS-CoV-2 attack rates; nevertheless, the role of chronic high-altitude exposure on the clinical outcome of critically ill COVID-19 patients has not been studied.

**Objective:** To compare the clinical course and outcomes of critically ill patients with COVID-19 hospitalized in two intensive care units (ICU) located at low and high altitude.

**Exposure and Outcome:** To explore the effect of two different elevations (10 m vs 2,850 m above sea level) on COVID-19 clinical outcome and survival.

**Methods:** A prospective cohort, two-center study in confirmed COVID-19 adult patients admitted to a low altitude (Sea level) and high altitude (2,850 m) ICU units in Ecuador was conducted. Two hundred and thirty confirmed COVID-19 patients were enrolled from March 15^th^ to July 15^th^, 2020. Sociodemographic, clinical, laboratory and imaging parameters including supportive therapies, pharmacological treatments and medical complications were reported and compared between the low and high-altitude groups.

**Results:** The median age of all the patients was 60 years, 64.8% were men and 35.2% were women. A total of 105 (45.7%) patients had at least one underlying comorbidity, the most frequent being chronic diseases, such as hypertension (33.5%), diabetes (16.5%), and chronic kidney failure (5.7%). The APACHE II scale at 72 hours was especially higher in the low-altitude group with a median of 18 points (IQR: 9.5-24.0), compared to 9 points (IQR: 5.0-22.0) obtained in the group of high altitude. There is evidence of a difference in survival in favor of the high-altitude group (p = 0.006), the median survival being 39 days, compared to 21 days in the low altitude group.

**Conclusion:** There has been a substantial improvement in survival amongst people admitted to the high-altitude critical care unit. High altitude living was associated with improved survival, especially among patients with no comorbidities. COVID-19 patients admitted to the high-altitude ICU unit have improved severity-of-disease classification system scores at 72 hours and reported better respiratory and ventilatory profiles than the low altitude group.

## Introduction

In December 2019, the first cases of pneumonia due to the SARS-CoV-2 virus were reported in Wuhan, China ^1,2^. In March 11^th^ 2020, the novel Coronavirus disease (COVID-19), a condition with multiple clinical features, which can rapidly evolve into acute respiratory distress syndrome (ARDS) and other serious complications was declared a pandemic ^2,3^. The disease spread rapidly, affecting regions and areas located in urban settings but also in rural and geographical distant areas all over the world^4,5^.

The epidemiological behavior of the pandemic showed an exponential growth throughout many countries, while others seem to have managed the outbreak better ^6^. Several studies have identified differences in morbidity and mortality depending on many factors including socioeconomic status, the burden of chronic diseases, adequate access to health care, the strength of the epidemiological surveillance systems and the implementation of control measures as well as strict mobility restrictions ^7^. During the first months of the pandemic, very few ecological studies showed a possible epidemiological and survival implication exerted by high altitude^7,8^. It has been proposed that these results are in part answered by the well-known physiological acclimatization and the long term adaptation to high altitude exposure, generating a greater tolerance to the hypoxia ^7–10^. These results are also supported by the presence of some biological plausibility regarding lower virus’ affinity for the type-2 angiotensin-converting enzyme (ACE2) receptor since highlanders might have lower expression of ACE-2 receptors^8^. Consequently, it has been hypothesized that high altitude populations would have less susceptibility to SARS-CoV-2, therefore lower likelihood to develop severe pathological conditions related to COVID-19 ^11^. Another hypothesis surrounding those findings refer to the involvement of the hypoxia-triggered protein that regulates angiogenesis, the well-known Hypoxia-inducible factor 1-alpha (HIF-1 α) ^10,12–16^.

The link between chronic exposure due to high altitude living and the clinical features of COVID-19 patients has been poorly studied^17,18^. Studies on clinical, ventilatory and respiratory support parameters’ differences have been studied in locations below 1,500 m and no evidence about the role of high-altitude exposure on severally ill COVID-19 patients have been published^19^.

Therefore, it is important to describe the possible role of high-altitude exposure as factor interfering with the clinical progression and the final outcome of patients with severe COVID-19.

## Methods

### Study design

A prospective cohort study including patients with severe SARS-CoV-2 infection confirmed with real-time polymerase chain reaction (RT-PCR) was performed from March 15^th^, 2020 to July 15^th^ 2020.

### Setting

The study was carried out in two intensive care units (ICU) located at two different elevations from the Social Security Health System (IESS) in Ecuador. The IESS-Quito Sur Hospital located in the city of Quito in Ecuador (located at 2,850 m above sea level) and the IESS-Los Ceibos located in the city of Guayaquil (Located at sea level).

### Population and study Size

All the patients included in this study were admitted to the ICU unit in one of the two hospitals. We used a non-probabilistic sampling technique for both hospitals based on the inclusion criteria. The present study included a total of 230 patients diagnosed with COVID-19 using the RT-PCR technique, of which 114 patients were treated in the high-altitude group (IESS-Quito Sur), while 116 patients belonged to the low-altitude group (IESS-Los Ceibos).

### Inclusion criteria

Adult men and women patients admitted to the ICU diagnosed with COVID-19 by means of RT-PCR who lived at least 1 year in the unit’s coverage region and signed the informed consent for the use of the information were included in this study.

### Exclusion criteria

Patients diagnosed with COVID-19 by RT-PCR who did not meet the criteria for admission to the ICU or who lived less than 1 year in the unit’s coverage region or who did not sign the informed consent for the use of the information were excluded.

### Data sources and variables

Demographic information, clinical characteristics (including medical history, history of symptoms, comorbidities), chest computed tomography (CT) results, laboratory findings, ventilatory values, and medications used were collected from each patient. The dates of disease onset, hospital admission, admission to the ICU, and death or discharge date from the ICU were also recorded, as well as the APACHE II scores and the Charlson index. The onset date was defined as the day the patient noticed any symptoms. The severity of COVID-19 was defined according to the diagnosis and treatment guide for SARS-CoV-2 issued by the World Health Organization (WHO) ^20^. It was designated as critical illness due to COVID-19 when patients had one of the following criteria: (a) acute respiratory distress syndrome (any grade); (b) Septicemia; and (c) septic shock. The data were obtained from the electronic medical record of a common registry system for both units and analyzed by three independent researchers.

### Statistical analysis

Categorical variables were summarized as frequencies and percentages, and continuous variables were described using median values and interquartile ranges (IQR) or mean and standard deviation, as appropriate. Analysis included two-tailed Student’s t-test and the Mann-Whitney U test was used. The frequencies of the categorical variables were compared using the chi-square test and expressed in count and percentage. Also, survival curves (Kaplan Meier), the log-rank statistic, and the hazard ratio between groups were obtained.

Bivariate and multivariate analyzes were performed to identify factors associated with death from COVID-19 in all patients using the Cox risk regression model. To obtain a reduced set of variables from the broad set of predictors, we carried out a progressive *en bloc* procedure assigning the predictor variables into six groups: sociodemographic characteristics and comorbidities, complications, scales, ventilatory values, medications, and laboratory and imaging parameters. A multivariate regression analysis was applied within each block using two criteria to achieve the best set of predictors: relevance to the clinical situation and bivariate and multivariate statistical significance (p <0.05). Variables with more than 25% missing values were not considered for the analysis.

All statistical analyzes were performed in SPSS version 25 and graphs were generated using GraphPad Prism version 7.00 software (GraphPad Software Inc).

### Bias

In order to minimize observation bias for systematic differences between the low and high-altitude group, observers were blinded the hypothesis under investigation and a strict protocol was implemented in both sites.

## Results

The present study included a total of n = 230 patients diagnosed with COVID-19, of which n = 114 patients were treated in the high-altitude group, while 116 patients belonged to the low-altitude group.

### Sociodemographic characteristic

The median age of all the patients was 60 years, with a range of 49 to 69 years, and the majority (80.9%) of them were over 45 years of age. More than half (64.8%) of the patients were men. The BMI of all the patients was 27.8 kg/m^2^, while about half (47.8%) were overweight, and (32.9%) some degree of obesity. A total of n = 105 (45.7%) patients had at least one underlying comorbidity, the most frequent being chronic diseases, such as hypertension (33.5%), diabetes (16.5%), and chronic kidney failure (5.7%). Five patients with COPD were identified (Table 1). When comparing the samples by altitude, no differences were evidenced between age, sex, and BMI. On the other hand, the low-altitude group presented a greater number of patients with comorbidities measured by the Charlson index, highlighting the cases of hypertensive and diabetic patients. The mean interval from the onset of symptoms to admission to the ICU for all patients was 8 days (IQR: 6-11). However, in the high-altitude group, there was a shorter median of 7 days (IQR: 5-10) (Table 1).

**Table 1.** Multivariate logistic regression analysis, regression coefficients and odds ratios (OR) calculated for demographic and independent risk factors in COVID-19 critically ill patients living at low and high altitude who were hospitalized in intensive care units

| **Category** | **All** | **High altitude** | **Low altitude** | **P-value** |
| --- | --- | --- | --- | --- |
|  | **n (%)** | **n (%)** | **n (%)** |  |
| Age - median (IQR) | 60.0 (49.0-69.0) | 55.5 (49.0-66.0) | 62.5 (48.5-69.0) | 0.181 |
| 24-34 | 13 (5.7) | 5 (4.4) | 8 (6.9) | 0.024 |
| 35-45 | 31 (13.5) | 15 (13.2) | 16 (13.8) | 0.024 |
| 46-56 | 58 (25.2) | 40 (35.1) | 18 (15.5) | 0.024 |
| 57-67 | 65 (28.3) | 26 (22.8) | 39 (33.6) | 0.024 |
| 68-78 | 54 (23.5) | 23 (20.2) | 31 (26.7) | 0.024 |
| Sex |  |  |  |  |
| Male | 149 (64.8) | 77 (67.5) | 72 (62.1) | 0.385 |
| Female | 81 (35.2) | 37 (32.5) | 44 (37.9) | 0.385 |
| BMI - median (IQR) | 27.8 (25.7-30.9) | 27.4 (24.9-31.0) | 28.2 (26.0-30.7) | 0.285 |
| Underweight | 1 (0.4) | 1 (0.9) | 0 (0.0) | 0.096 |
| Normal weight | 43 (18.9) | 27 (24.1) | 16 (13.8) | 0.096 |
| Overweight | 109 (47.8) | 48 (42.9) | 61 (52.6) | 0.096 |
| Obesity class I | 59 (25.9) | 25 (22.3) | 34 (29.3) | 0.096 |
| Obesity class II | 15 (6.6) | 10 (8.9) | 5 (4.3) | 0.096 |
| Obesity class III | 1 (0.4) | 1 (0.9) | 0 (0.0) | 0.096 |
| Charlson index |  |  |  |  |
| Presence of |  |  |  |  |
| Comorbidities | 135 (58.7) | 85 (74.6) | 50 (43.1) | 0.000 |
| Low comorbidity | 28 (12.2) | 10 (8.8) | 18 (15.5) | 0.000 |
| Hight comorbidity | 67 (29.1) | 19 (16.7) | 48 (41.4) | 0.000 |
| Comorbidity | 105 (45.7) | 33 (28.9) | 72 (62.1) | 0.000 |
| Arterial Hypertension | 77 (33.5) | 21 (18.4) | 56 (48.3) | 0.000 |
| CKD | 13 (5.7) | 3 (2.6) | 10 (8.6) | 0.049 |
| Asthma | 2 (0.9) | 1 (0.9) | 1 (0.9) | 0.99 |
| Diabetes | 38 (16.5) | 12 (10.5) | 26 (22.4) | 0.015 |
| Psoriasis | 2 (0.9) | 0 (0.0) | 2 (1.7) | 0.159 |
| CHF | 2 (0.9) | 1 (0.9) | 1 (0.9) | 0.99 |
| Cancer | 4 (1.7) | 2 (1.8) | 2 (1.7) | 0.986 |
| CVA | 2 (0.9) | 0 (0.0) | 2 (1.7) | 0.159 |
| COPD | 5 (2.2) | 4 (3.5) | 1 (0.9) | 0.169 |
| Rheumatoid arthritis | 2 (0.9) | 0 (0.0) | 2 (1.7) | 0.159 |
| TB | 2 (0.9) | 2 (1.8) | 0 (0.0) | 0.152 |
| Hepatic cirrhosis | 2 (0.9) | 1 (0.9) | 1 (0.9) | 0.99 |
| Pulmonary fibrosis | 3 (1.3) | 1 (0.9) | 2 (1.7) | 0.571 |
| Hypothyroidism | 5 (2.2) | 3 (2.6) | 2 (1.7) | 0.637 |

Abbreviations: IQR: Interquartile range; BMI: body mass index; CKD: Chronic kidney disease; CHF: Chronic hepatic failure; CVD: Cerebro-vascular Disease; COPD: Chronic obstructive pulmonary disease; TB: Tuberculosis

Regarding the scales evaluated, it was evidenced that upon admission to the ICU, the APACHE II scale in the first 24 hours presented a median of 15 points (IQR: 10.0-20.0) in the high-altitude group, whilst the low altitude scored 16 points (12.0-20.5) and did not show a statistical difference for both groups (p = 0.206). However, the same scale at 72 hours was especially higher in the low-altitude group with a median of 18 points (IQR: 9.5-24.0), compared to 9 points (IQR: 5.0-22.0) obtained in the group of high altitude. Concerning the most common complications presented during the ICU stay, acute respiratory distress syndrome in adults was evidenced in n = 219 (95.2%) patients, any type of shock in n = 166 (72.2%) patients, acute / exacerbated renal failure in n = 101 (43.9%) and delirium in n = 88 (38.3%). There were no statistical relationships between complications and altitude (Table 2). Finally, n = 129 deaths (56.1%) were recorded in the entire sample, of which most (n =77 (66.4%) were recorded in the low-altitude group compared to n = 52 (45.6%) at high altitude, p = 0.002 (Table 2).

**Table 2.**
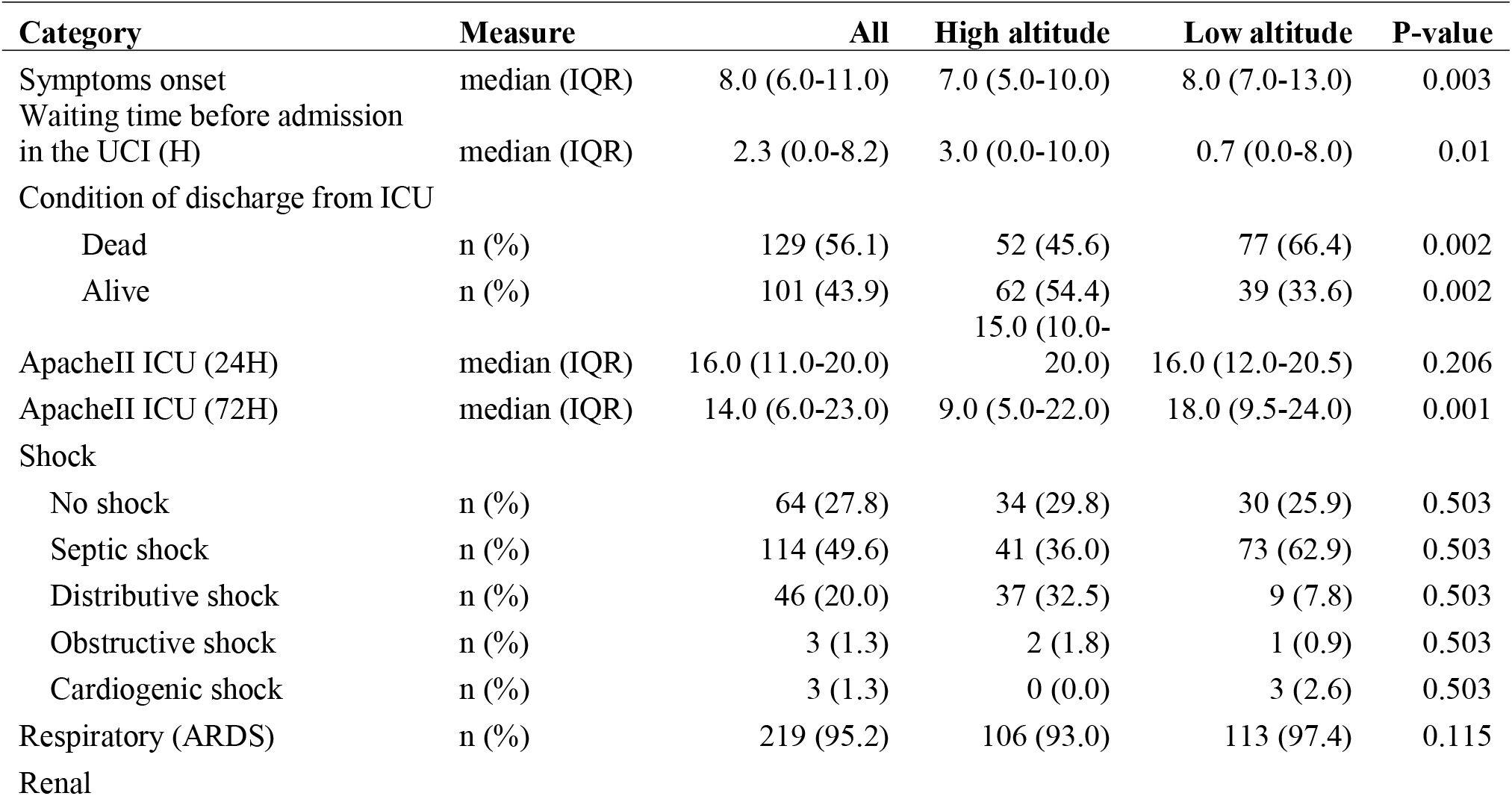
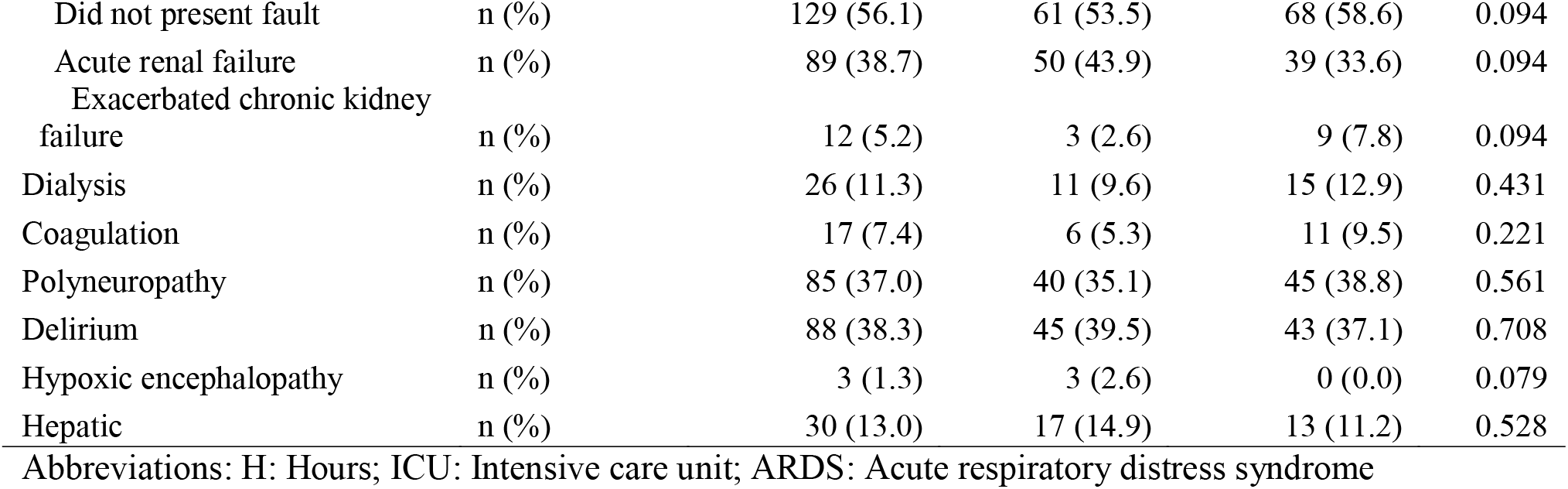
Table 1 Multivariate logistic regression analysis, regression coefficients and odds ratios (OR) calculated for clinical predictors for COVID-19 mortality among critically ill patients living at low and high-altitude who were hospitalized in intensive care units

The laboratory results are shown in (Table 3), where lower values of platelets, liver enzymes (AST and ALT), and lactate were evidenced in the low altitude group compared to the high-altitude group. Against the leukocyte count was higher in the low altitude group (Figure 1).

**Table 3.** Analysis of the mean and median differences of the principal hematological and serological parameters in severely ill patients with covid19

| Category | All | High altitude | Low altitude | P value |
| --- | --- | --- | --- | --- |
|  | median (IQR) | median (IQR) | median (IQR) |  |
| **Hematic Biometry** |  |  |  |  |
| Hemoglobin mg/dL | 13.7 (12.2-14.7) | 13.7 (12.4-14.8) | 13.5 (11.8-14.6) | 0.095 |
| Hemoglobin mg/dL (72H) | 12.3 (11.1-13.6) | 12.4 (11.5-13.6) | 12.1 (10.9-13.6) | 0.268 |
| Hemoglobin mg/dL (7D) | 11.6 (10.0-12.9) | 11.8 (10.3-13.0) | 11.1 (9.9-12.8) | 0.214 |
| Leukocytes 10<sup>3</sup>/μL | 11.8 (8.9-16.1) | 10.5 (8.2-14.4) | 13.0 (10.2-17.4) | 0.000 |
| Leukocytes 10<sup>3</sup>/μL (72) | 11.3 (8.7-14.2) | 10.0 (8.3-12.4) | 12.5 (9.4-16.6) | 0.000 |
| Leukocytes 10<sup>3</sup>/μL (7D) | 11.8 (9.3-16.9) | 10.8 (8.7-12.7) | 13.5 (10.2-20.7) | 0.000 |
| Neutrophils % | 86.7 (81.0-90.0) | 87.0 (81.0-90.0) | 86.2 (81.3-89.5) | 0.215 |
| Neutrophils % (72H) | 87.0 (82.0-90.0) | 87.0 (82.1-90.0) | 87.0 (81.0-90.0) | 0.723 |
| Neutrophils % (7D) | 85.0 (78.9-89.0) | 83.5 (79.6-88.0) | 86.5 (77.6-90.0) | 0.406 |
| Lymphocytes 10<sup>3</sup>/μL | 6.7 (4.2-10.2) | 6.7 (4.4-10.0) | 6.1 (4.0-10.9) | 0.994 |
| Lymphocytes 10<sup>3</sup>/μL (72H) | 6.0 (4.0-9.3) | 6.0 (4.0-8.7) | 6.0 (4.0-9.8) | 0.844 |
| Lymphocytes 10<sup>3</sup>/μL (7D) | 7.3 (4.0-11.4) | 8.0 (4.7-10.9) | 6.0 (4.0-12.0) | 0.463 |
| Platelets 10<sup>3</sup>/μL | 290.0 (215.0-378.0) | 277.0 (218.0-367.0) | 300.5 (213.0-387.0) | 0.605 |
| Platelets 10<sup>3</sup>/μL (72H) | 283.0 (202.0-369.0) | 300.5 (228.0-383.0) | 255.5 (185.0-347.0) | 0.015 |
| Platelets 10<sup>3</sup>/μL (7D) | 262.0 (185.0-401.0) | 306.0 (207.0-435.0) | 241.0 (152.0-338.0) | 0.002 |
| **Blood chemistry** |  |  |  |  |
| D-Dimer ng/ml | 2,193.0 (796.0-6,293.0) | 1,895.5 (743.0-4,745.0) | 2,600.0 (800.0-7,700.0) | 0.475 |
| D-Dimer ng/ml (72H) | 2,003.0 (738.0-5,400.0) | 2,347.0 (947.0-3,800.0) | 1,600.0 (450.0-6,700.0) | 0.745 |
| Urea mg/dL | 42.0 (27.0-64.0) | 36.4 (23.5-57.7) | 45.5 (30.0-66.5) | 0.011 |
| Urea mg/dL (72H) | 53.0 (36.8-83.5) | 51.0 (36.4-83.0) | 54.5 (37.0-95.0) | 0.566 |
| Urea mg/dL (7D) | 53.0 (36.0-92.0) | 51.3 (38.3-85.3) | 54.5 (33.0-100.0) | 0.839 |
| Creatinine mg/dL | 0.8 (0.7-1.2) | 0.8 (0.7-1.1) | 0.8 (0.7-1.3) | 0.150 |
| Creatinine mg/dL (72H) | 0.8 (0.6-1.6) | 0.8 (0.7-1.7) | 0.8 (0.6-1.5) | 0.298 |
| Creatinine mg/dL (7D) | 0.8 (0.6-1.6) | 0.8 (0.6-1.3) | 0.8 (0.6-2.0) | 0.982 |
| ALT U/L | 47.0 (30.0-78.0) | 52.0 (33.5-79.0) | 42.0 (27.0-75.0) | 0.080 |
| ALT U/L (72) | 44.0 (28.0-64.0) | 49.0 (40.0-67.5) | 40.0 (27.0-60.0) | 0.027 |
| ALT U/L (7D) | 43.0 (30.0-67.0) | 55.0 (38.0-86.0) | 39.0 (24.0-65.0) | 0.006 |
| AST U/L | 48.0 (31.0-62.0) | 49.0 (34.5-67.5) | 45.0 (31.0-59.0) | 0.137 |
| AST U/L (72) | 38.0 (26.0-60.0) | 47.0 (34.0-63.5) | 34.0 (23.0-57.0) | 0.002 |
| AST U/L (7D) | 40.0 (28.0-57.0) | 47.0 (35.0-66.0) | 34.0 (27.0-56.0) | 0.024 |
| Ferritin µg/ml | 1,469.0 (912.2-2,000.0) | 1,418.8 (824.0-1,979.0) | 1,600.0 (1,047.0-2,000.0) | 0.300 |
| Ferritin µg/ml (72H) | 1,670.5 (1,006.5-2,000.0) | 1,615.5 (1,286.0-2,000.0) | 1,670.5 (872.5-2,000.0) | 0.497 |
| LDH U/L | 402.5 (315.0-584.0) | 382.0 (303.0-541.0) | 427.0 (320.0-608.0) | 0.134 |
| LDH U/L (72H) | 353.0 (253.0-459.0) | 353.0 (251.0-459.0) | 356.5 (261.0-515.5) | 0.559 |
| LDH U/L (7D) | 267.0 (180.0-365.0) | 205.0 (167.0-355.0) | 293.0 (180.0-377.0) | 0.431 |
| CRP mg/L | 27.0 (18.5-45.0) | 22.8 (18.1-33.3) | 36.0 (19.0-48.0) | 0.004 |
| CRP mg/L (72H) | 22.6 (5.9-37.6) | 22.1 (14.6-32.0) | 23.2 (2.2-44.0) | 0.809 |
| CRP mg/L (7D) | 15.0 (2.5-36.1) | 20.8 (14.6-29.7) | 11.5 (1.8-38.0) | 0.209 |
| PCT ng/ml | 0.5 (0.2-1.8) | 0.5 (0.2-1.9) | 0.6 (0.2-1.6) | 0.953 |
| PCT ng/ml (72H) | 0.8 (0.2-2.8) | 0.7 (0.2-1.0) | 1.0 (0.3-3.1) | 0.531 |
| PCT ng/ml (7D) | 0.6 (0.2-2.3) | 0.2 (0.2-1.2) | 0.6 (0.1-2.6) | 0.370 |
| Gasometry |  |  |  |  |
| ph | 7.4 (7.3-7.4) | 7.4 (7.3-7.5) | 7.3 (7.2-7.4) | 0.000 |
| ph (72H) | 7.4 (7.3-7.4) | 7.4 (7.3-7.4) | 7.4 (7.2-7.4) | 0.003 |
| ph (7D) | 7.4 (7.3-7.5) | 7.4 (7.3-7.5) | 7.4 (7.3-7.4) | 0.001 |
| SaO<sub>2</sub> % | 94.0 (89.0-97.0) | 91.0 (87.0-94.0) | 96.9 (93.0-98.0) | 0.000 |
| SaO<sub>2</sub> % (72H) | 96.0 (93.0-98.0) | 94.0 (92.0-95.4) | 97.8 (96.0-98.5) | 0.000 |
| SaO<sub>2</sub> % (7D) | 96.0 (92.0-98.0) | 94.0 (91.0-96.0) | 97.7 (96.0-98.2) | 0.000 |
| PaO<sub>2</sub> mmHg | 76.0 (59.0-104.0) | 65.0 (52.0-76.0) | 97.9 (72.3-138.4) | 0.000 |
| PaO<sub>2</sub> mmHg (72H) | 88.3 (70.0-124.3) | 72.8 (66.0-83.0) | 115.5 (92.0-150.0) | 0.000 |
| PaO<sub>2</sub> mmHg (7D) | 82.5 (67.0-119.8) | 69.5 (61.3-80.7) | 110.5 (88.0-145.0) | 0.000 |
| PaCO<sub>2</sub> mmHg | 38.0 (31.7-48.3) | 35.8 (31.0-43.0) | 43.0 (32.5-56.3) | 0.007 |
| PaCO<sub>2</sub> mmHg (72H) | 43.1 (37.0-53.9) | 42.0 (37.0-50.0) | 45.3 (37.0-57.6) | 0.212 |
| PaCO<sub>2</sub> mmHg (7D) | 42.0 (38.0-51.0) | 42.7 (38.0-52.0) | 41.9 (37.2-50.2) | 0.774 |
| HCO<sub>3</sub> mEq/L | 20.9 (18.7-23.6) | 21.9 (19.1-24.9) | 20.2 (17.7-22.6) | 0.000 |
| HCO<sub>3</sub> mEq/L (72H)-mean (SD) | 23.46 (5.70) | 24.59 (5.85) | 22.36 (5.35) | 0.003 |
| HCO<sub>3</sub> mEq/L (7D) | 25.3 (21.5-29.1) | 28.0 (22.1-30.6) | 24.2 (20.0-26.9) | 0.000 |
| Lactate mmol/L | 1.8 (1.3-2.3) | 1.8 (1.4-2.3) | 1.7 (1.0-2.1) | 0.800 |
| Lactate mmol/L (72H) | 1.6 (1.2-2.1) | 1.7 (1.3-2.2) | 1.4 (1.0-2.0) | 0.820 |
| Lactate mmol/L (7D) | 1.6 (1.2-2.0) | 1.7 (1.3-2.1) | 1.4 (1.0-2.0) | 0.100 |

Abbreviations: H: Hours; D: Days; ALT: Alanine aminotransferase; AST: Aspartate aminotransferase; LDH: lactate dehydrogenase; PCR: C-reactive protein; PCT: procalcitonin; ph: Potential of hydrogen; SaO<sub>2</sub>: Oxygen saturation of arterial blood; PaO<sub>2</sub>: Partial pressure of oxygen in arterial blood; PaCO<sub>2</sub>: Partial pressure of carbon dioxide in arterial blood; HCO<sub>3</sub>: Serum bicarbonate.

**Figure 1.**
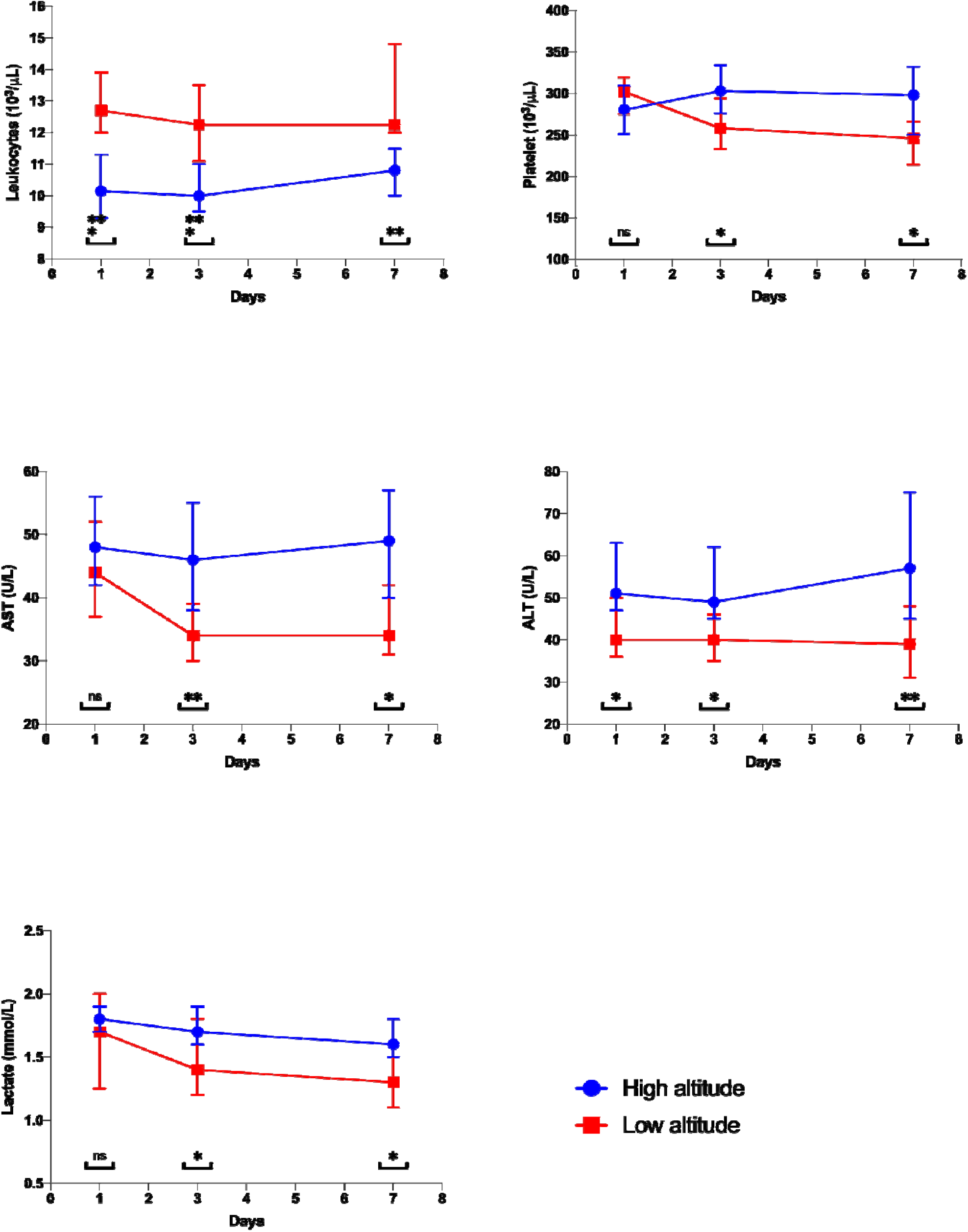
Hematological differences in the low and high-altitude group. Abbreviations: ALT: Alanine aminotransferase; AST: Aspartate aminotransferase.

The acid-base profile for both groups show normal ranges; however, there is a greater alkalotic component (higher median pH and bicarbonate) in the low-altitude group (p=0.000). While the level of CO_2_ appears in normal ranges and without difference between the two groups (Table 3).

Mechanical ventilation was maintained for a median of 12 days (IQR: 7.0-20.0) for the two groups. During admission, the FiO_2_ for both groups did not show differences, however, the measurements at 72 hours and 7 days show significant differences with higher values in the low altitude group (Figure 2).

**Figure 2.**
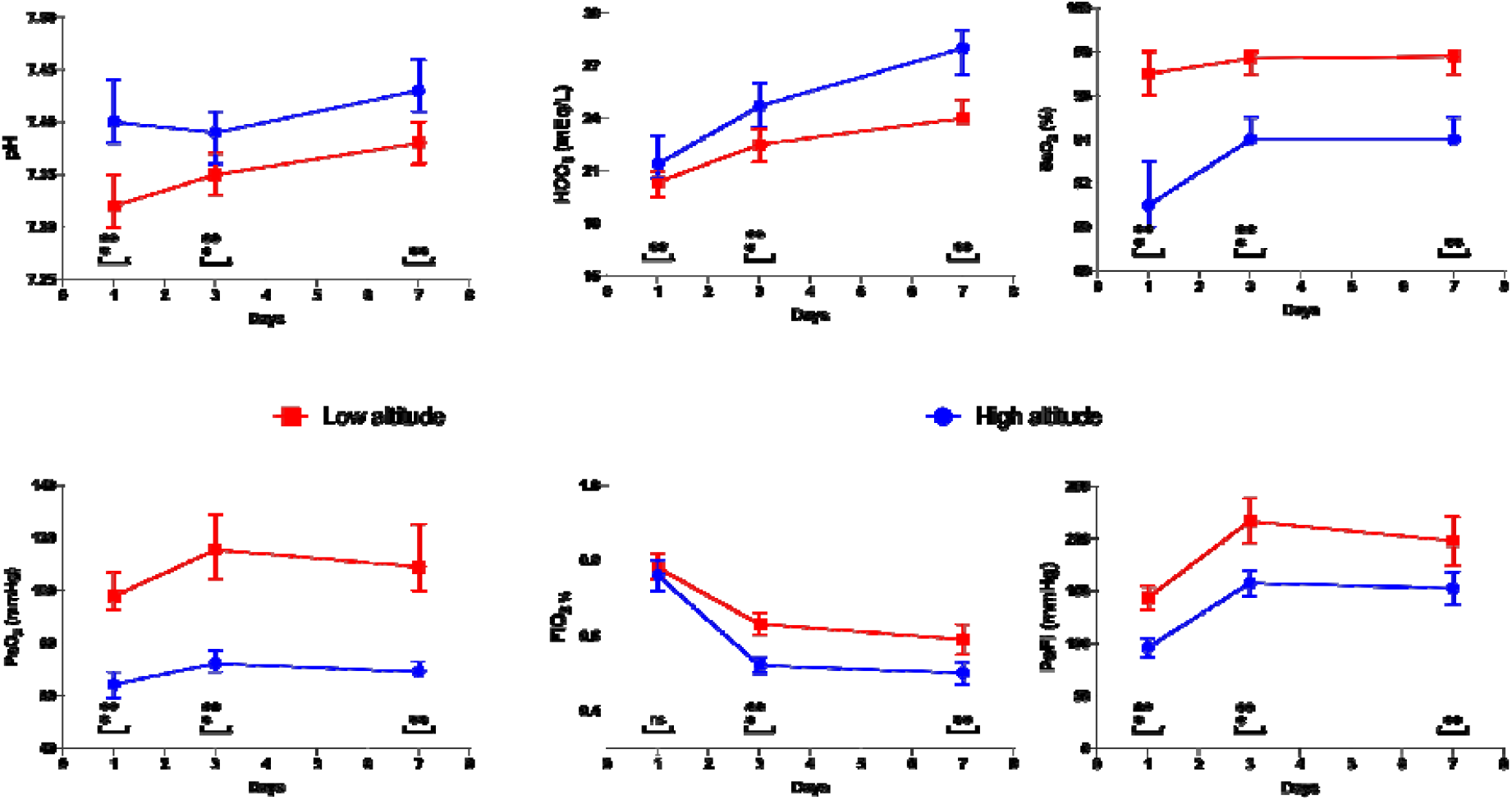
Respiratory differences

Similarly, a higher value was evidenced during the measurements of the partial pressure of oxygen (PO_2_) in the patients in the low altitude group, compared to the high-altitude group and consequently in the Pa-Fi relationship (Figure 2). The need for a tracheostomy reached 17.5% of patients (Table 4).

**Table 4.** Ventilatory and pulmonary parameters among COVID-19 patients in the low and high-altitude group

| Category | Measure | All | High altitude | Low altitude | P-value |
| --- | --- | --- | --- | --- | --- |
| Received mechanical ventilation | n (%) | 204 (88.7) | 105 (92.1) | 99 (85.3) | 0.105 |
| No Invasive | n (%) | 26 (11.3) | 12 (10.5) | 14 (12.1) | 0.712 |
| Control Pressure | n (%) | 121 (52.6) | 96 (84.2) | 25 (21.6) | 0.000 |
| Support Pressure | n (%) | 13 (5.7) | 12 (10.5) | 1 (0.9) | 0.002 |
| Control volume | n (%) | 134 (58.3) | 36 (31.6) | 98 (84.5) | 0.000 |
| High Flow | n (%) | 12 (5.2) | 0 (0.0) | 12 (10.3) | 0.000 |
| Airway Pressure release ventilation | n (%) | 5 (2.2) | 5 (4.4) | 0 (0.0) | 0.023 |
| Recruitment | n (%) | 72 (31.3) | 33 (28.9) | 39 (33.6) | 0.445 |
| Tracheostomy | n (%) | 43 (18.7) | 23 (20.2) | 20 (17.2) | 0.568 |
| Pronation |  |  |  |  |  |
| Intermittent | n (%) | 56 (24.3) | 18 (15.8) | 38 (32.8) | 0.014 |
| Extended | n (%) | 126 (54.8) | 68 (59.6) | 58 (50.0) | 0.014 |
| Pronation (H) | median (IQR) | 50.0 (16.0-96.0) | 48.0 (2.0-72.0) | 72.0 (24.0-120.0) | 0.000 |
| Volume | median<br>(IQR) | 390.0 (360.0-420.0) | 400.0 (380.0-420.0) | 380.0 (360.0-400.0) | 0.000 |
| Volume (72H) | median<br>(IQR) | 400.0 (360.0-420.0) | 418.0 (380.0-450.0) | 380.0 (360.0-400.0) | 0.000 |
| RR | median<br>(IQR) | 22.0 (20.0-24.0) | 20.0 (20.0-22.0) | 22.0 (20.0-25.0) | 0.000 |
| RR (72H) | median<br>(IQR) | 22.0 (20.0-24.0) | 22.0 (20.0-24.0) | 22.0 (22.0-24.0) | 0.020 |
| FiO<sub>2</sub> | median<br>(IQR) | 0.8 (0.6-1.0) | 0.7 (0.6-1.0) | 0.8 (0.6-1.0) | 0.000 |
| FiO<sub>2</sub> (72H) | median<br>(IQR) | 0.6 (0.5-0.7) | 0.5 (0.5-0.6) | 0.6 (0.6-0.7) | 0.020 |
| FiO<sub>2</sub> (7D) | median<br>(IQR) | 0.5 (0.4-0.7) | 0.5 (0.4-0.6) | 0.6 (0.5-0.8) | 0.000 |
| Oxygen ml kg | mean (SD) | 6.60 (0.91) | 6.95 (0.94) | 6.23 (0.73) | 0.000 |
| Oxygen ml kg (72H) | mean (SD) | 6.64 (0.99) | 7.03 (1.06) | 6.23 (0.69) | 0.000 |
| PAFI mmHg | median<br>(IQR) | 104.3 (72.9-154.5) | 87.0 (61.0-121.0) | 133.1 (94.4-176.6) | 0.000 |
| PAFI mmHg (72H) | median<br>(IQR) | 165.0 (128.3-216.7) | 150.0 (125.5-192.4) | 187.5 (147.2-255.0) | 0.000 |
| PAFI mmHg (7D) | median<br>(IQR) | 162.5 (122.0-231.0) | 149.6 (119.0-184.8) | 200.0 (125.0-263.6) | 0.001 |
| PEEP | median<br>(IQR) | 12.0 (10.0-14.0) | 12.0 (10.0-14.0) | 10.0 (9.0-12.0) | 0.000 |
| PEEP (72H) | median<br>(IQR) | 10.0 (9.0-12.0) | 12.0 (9.0-14.0) | 10.0 (8.0-12.0) | 0.002 |
| Peak Pressure | median<br>(IQR) | 30.0 (28.0-32.0) | 28.0 (25.0-31.0) | 31.0 (29.0-35.0) | 0.000 |
| Peak Pressure (72H) | median<br>(IQR) | 30.0 (26.0-32.0) | 28.0 (24.0-30.0) | 31.0 (28.0-33.0) | 0.000 |
| Plateau Pressure | median<br>(IQR) | 26.0 (23.0-28.0) | 25.0 (22.0-28.0) | 26.0 (24.0-29.0) | 0.047 |
| Plateau Pressure (72H) | median<br>(IQR) | 25.0 (21.0-27.0) | 24.0 (20.0-27.0) | 25.5 (22.0-28.0) | 0.029 |
| Static Compliance | median<br>(IQR) | 28.0 (22.0-37.0) | 30.5 (26.0-37.0) | 27.0 (18.0-34.0) | 0.006 |
| Static Compliance (72H) | mean (SD) | 31.65 (10.92) | 33.63 (9.48) | 29.53 (11.97) | 0.008 |
| Resistors | median<br>(IQR) | 10.0 (8.0-15.0) | 11.0 (9.0-14.0) | 10.0 (3.0-15.0) | 0.378 |
| Resistors | median<br>(IQR) | 10.0 (7.0-12.0) | 10.0 (9.0-12.0) | 10.0 (0.0-12.5) | 0.023 |
| Driving Pressure (24H) | median<br>(IQR) | 13.0 (9.0-16.0) | 12.0 (9.0-15.0) | 14.0 (8.0-18.0) | 0.022 |
| Driving Pressure (72H) | median<br>(IQR) | 12.5 (9.0-16.0) | 12.0 (8.0-14.0) | 14.0 (9.0-17.0) | 0.021 |
| Mechanic power | median<br>(IQR) | 18.0 (14.0-21.3) | 17.2 (13.2-20.6) | 18.9 (14.7-22.5) | 0.073 |
| Mechanic power (72H) | median<br>(IQR) | 18.4 (13.5-22.2) | 18.0 (13.2-22.2) | 18.7 (14.2-22.1) | 0.907 |

Abbreviations: H: Hours; D: Days; FiO<sub>2</sub>: Fraction of inspired oxygen; RR:Respiratory rate ; PAFI: Relationship between the alveolar-arterial oxygen gradient and PaO<sub>2</sub>/FiO<sub>2</sub>; PEEP: Positive End-Expiratory Pressure

### Medicines

During the hospital stay, 136 (63.6%) patients received corticosteroids, of which up to 60 (39%) in mg/kg doses, for a median time of 4 (IQR: 3-6) days. The most prescribed corticosteroid was methylprednisolone (n = 97; 42.2%). A total of n = 224 (97.4%) of patients received heparins, of which 82 (79.1%) patients received isocoagulation doses. Regarding medications that to date were used for the treatment of COVID, it was evidenced that 118 (51.3%) and 149 (64.8%) of patients received Hydroxychloroquine and Lopinavir / Ritonavir, respectively. In the comparison between groups, the low-altitude group received greater numbers of corticosteroid prescriptions and tocilizumab than the high-altitude group. However, the patients did not show differences in the administration of heparins, antimalarials, or lopinavir/ritonavir (Table 5).

**Table 5.** Pharmaceutical treatment in the low and high-altitude group

| Category | All | High altitude | Low altitude | P-value |
| --- | --- | --- | --- | --- |
| Heparins | 224 (97.4) | 71 (98.3) | 112 (96.6) | 0.420 |
| Heparin at isocoagulation doses | 182 (79.1) | 71 (62.3) | 111 (95.7) | 0.000 |
| Heparin at anticoagulation doses | 42 (18.3) | 41 (36.0) | 1 (0.9) | 0.000 |
| Corticosteroids | 146 (63.6) | 59 (51.8) | 87 (75.0) | 0.000 |
| Methylprednisolone | 97 (42.2) | 40 (35.1) | 57 (49.1) | 0.000 |
| Dexamethasone | 34 (14.8) | 5 (4.4) | 29 (25.0) | 0.000 |
| Hydrocortisone | 2 (0.9) | 2 (1.8) | 0 (0.0) | 0.000 |
| Prednisone | 13 (5.7) | 12 (10.5) | 1 (0.9) | 0.000 |
| Corticosteroids Days - median (IQR) | 4.0 (3.0-6.0) | 3.0 (3.0-3.0) | 5.0 (3.0-7.0) | 0.000 |
| Corticosteroids Doses |  |  |  |  |
| mg/kg | 60 (39.0) | 26 (43.3) | 34 (36.2) | 0.649 |
| Lower Doses | 48 (31.2) | 18 (30.0) | 30 (31.9) | 0.649 |
| Pulses | 46 (29.9) | 16 (26.7) | 30 (31.9) | 0.649 |
| Antimalarials | 168 (73.0) | 88 (77.2) | 80 (69.0) | 0.312 |
| Hydroxychloroquine | 50 (21.7) | 13 (11.4) | 37 (31.9) | 0.000 |
| Chloroquine | 118 (51.3) | 75 (65.8) | 43 (37.1) | 0.000 |
| Others |  |  |  |  |
| Lopinavir/Ritonavir | 149 (64.8) | 69 (60.5) | 80 (69.0) | 0.180 |
| Tocilizumab | 23 (10.0) | 3 (2.6) | 20 (17.2) | 0.000 |

### Altitude and mortality from COVID

There is evidence of a difference in survival in favor of the high-altitude group (p = 0.006), with the median survival of 39 days, compared to 21 days of the low altitude group (Figure 3).

**Figure 3.**
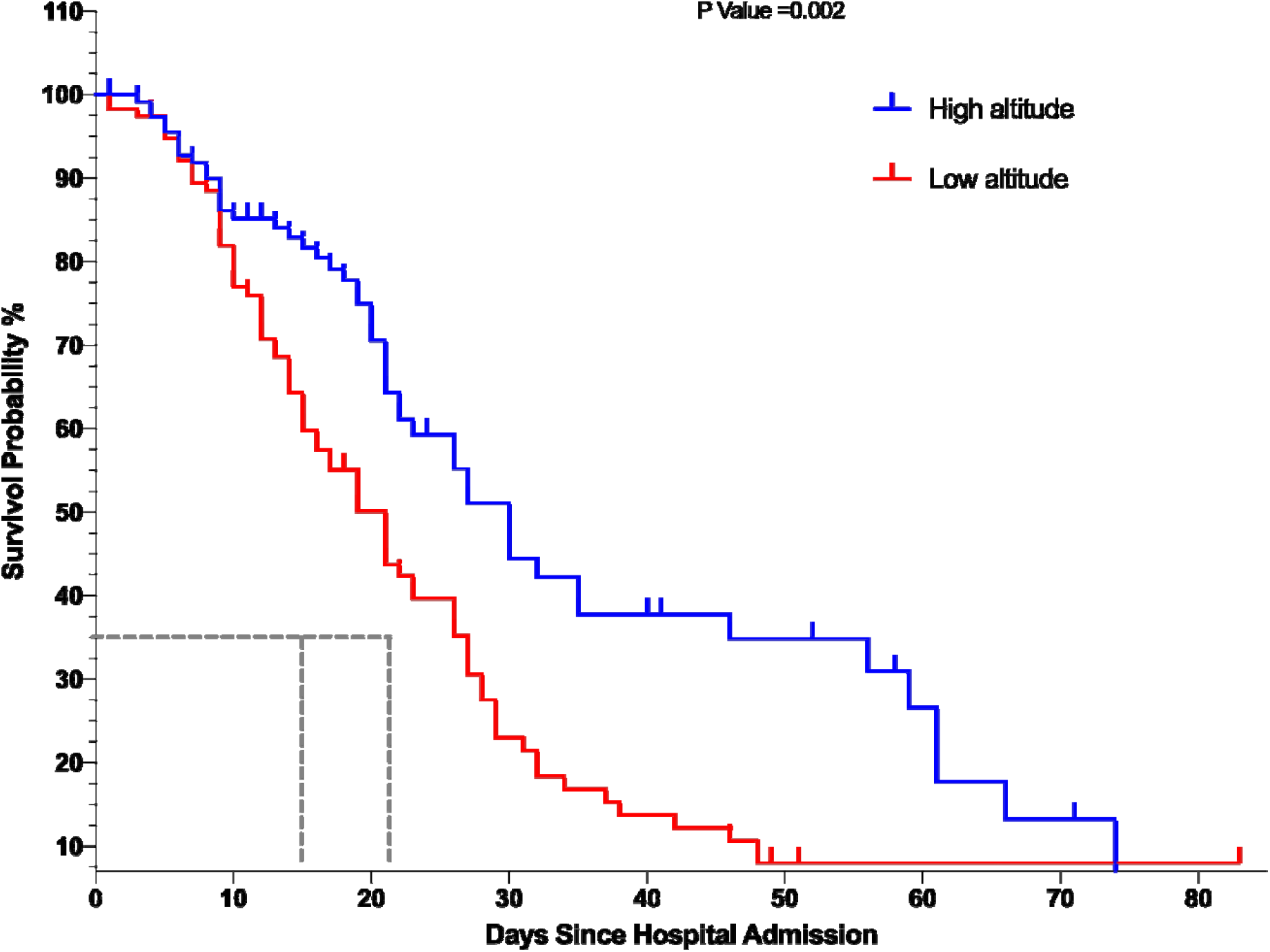
The Kaplan-Meier curves for mortality according to altitude

The hazard ratio found was 0.55 (95%CI = 0.39-0.78). Due to the differences found concerning comorbidities, the analysis by subgroups was performed according to the Charlson classification (Figure 4).

**Figure 4.**
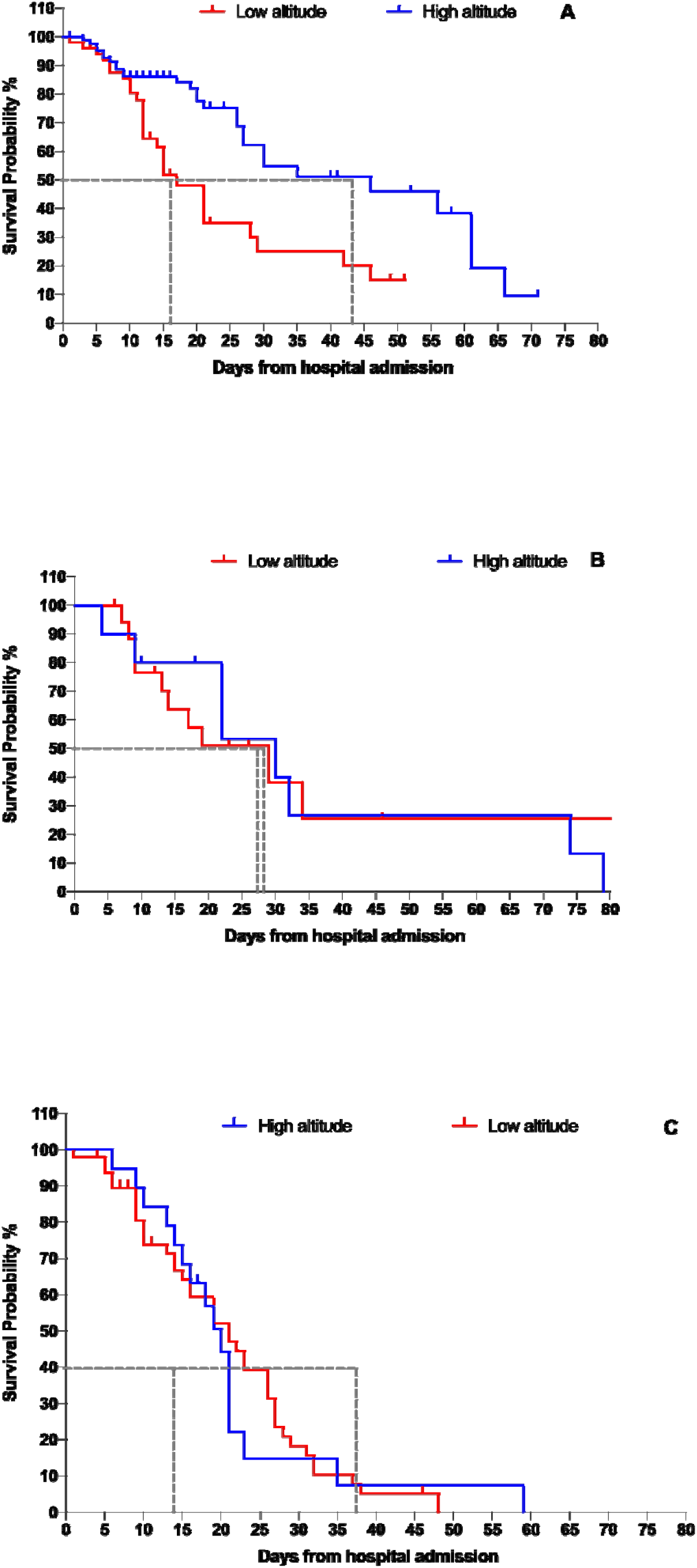
The Kaplan-Meier curves for mortality according to altitude. 4A Curve with more comorbidtiesties p=0.920 4B p:0.929 Curve with fewer comorbidities 4C Curve with no comorbidities P Value: 0.005

It is evident that the patients classified with a low and high index did not present differences between groups by altitude p = 0.929 and p = 0.920, respectively. However, in the group with no comorbidities, there was evidence of a difference between altitudes (p = 0.005), with the median survival of 17 days in the low-altitude group and 49 in the high-altitude group. The hazard ratio found was 0.41 (95% CI = 0.23-0.75).

### Predictors of death

In the final adjusted analysis, 5 factors associated with the risk of death were found. As a protection factor, they were high altitude and the presence of a tracheostomy. On the other hand, as risk factors were the APACHE II score greater than 17 at 72 hours, the relationship between arterial oxygen pressure and inspiratory oxygen fraction (PaO_2_ / FiO_2_) on the seventh day less than 300, and the presence of coagulopathy during the hospitalization (Figure 5).

**Figure 5.**
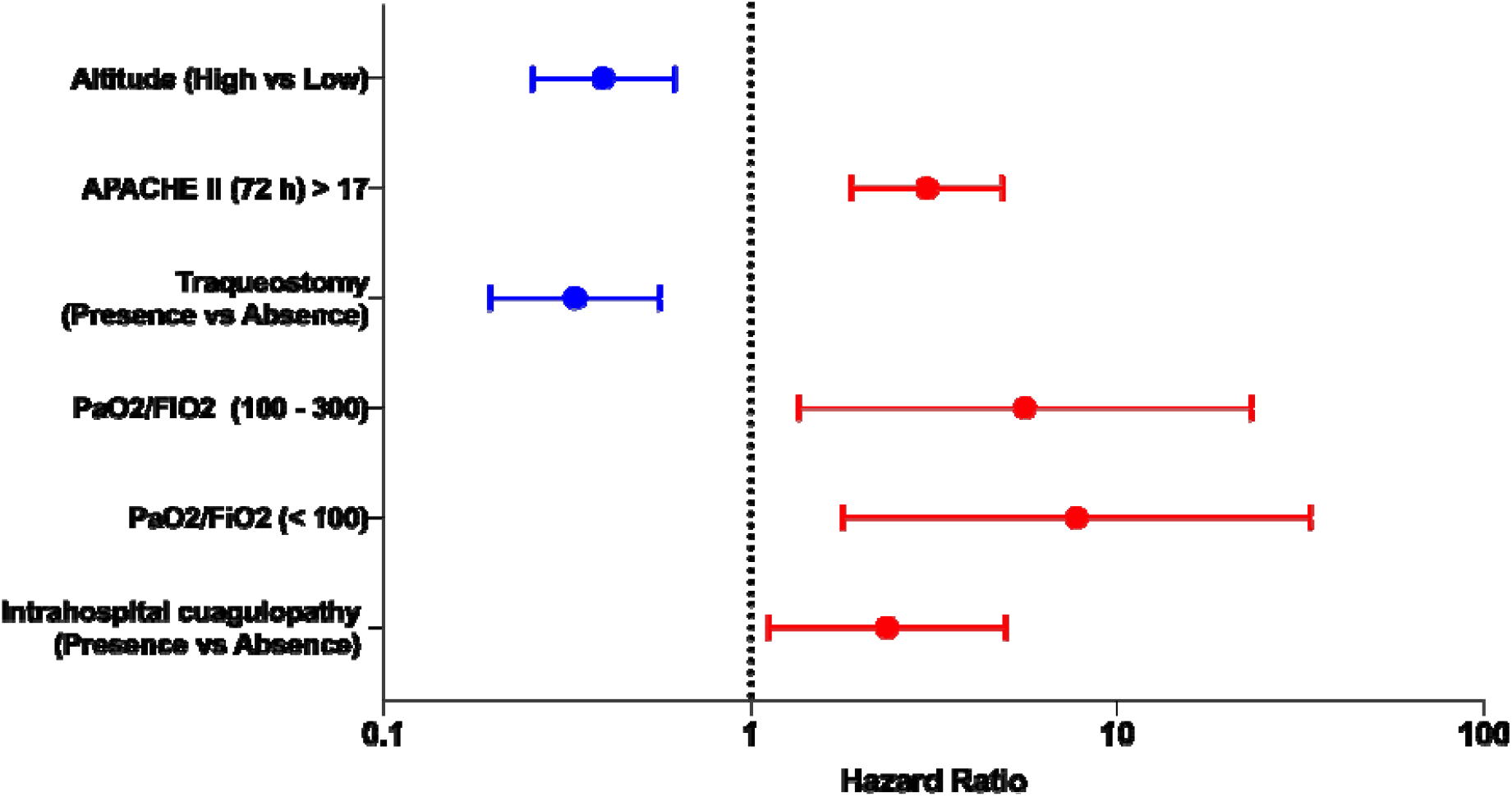
High altitude mortality predictorsCaracterística. High Altitude vs Low Altitude HR 0.40 (CI 95% 0.25 - 0.62), Intra Hospital Cuagulopathy (Presence vs Abcense HR 2.35 (CI 95% 1.12 - 4.95), PaO2/FiO2 <100 HR 7.74 (CI 95% 1.79 - 33.48), PaO2/FiO2 100 – 300 HR 5.60 (CI 95% 1.36 - 23.14), Traqueostomy (presence vs absence HR 0.33 (CI 95% 0.20 - 0.56), APACHE II (72 hrs) > 17 HR 3.03 (CI 95% 1.88 - 4.87).

## Discussion

The COVID-19 pandemic has severely affected several countries worldwide, so understanding the factors involved in its prognosis is a priority worldwide. Evidence has been found of the influence of altitude on the number of infections and mortality, with lower values in high altitude places ^21,22^. The pathophysiological reason for this relationship is not established. On the one hand, high-altitude patients present a chronic conversion of the hypoxia-inducing factor type 1 to type 2, which favors a greater tolerance to hypoxemia and decreases the acute tissue damage triggered by patients with severe acute respiratory conditions. On the other hand, it has been shown that some high-altitude resident population groups have developed polymorphisms of the ACE2 receptor that favor a better tolerance to hypoxia ^23,24^. The same receptor that is used by SARS-CoV-2 to enter the target cells and that, when inactivated, promotes a pro-inflammatory state that would increase the repercussions in the lungs and other organs ^11,23,24^. Despite the absence of clear pathophysiology, the present study provides relevant epidemiological and clinical data for understanding the influence of altitude on the evolution of seriously ill patients with COVID-19.

The study shows a mainly male hospital population with a median age of 60 years, with results similar to other studies ^3,25–27^. The analysis together showed leukocytosis with neutrophilia and lymphopenia, besides, increased values of ferritin, LDH, CRP and procalcitonin were evidenced. This analysis is compatible with studies similar to those found in patients with severe COVID-19, where abnormal values of leukocytes, neutrophils, lymphocytes and platelets are evidenced ^28,29^, while blood biochemistry shows increased CRP and LDH ^30^. In the analysis by altitude, the low altitude group had a higher number of leukocytes, a lower number of platelets and higher levels of CRP, however the rest of the values were similar in both groups.

The oxygenation parameters expressed in levels of PaO_2_ / FiO_2_, PO_2_ and SaO_2_ were found to be compromised in both groups at the time of admission. Findings similar to those found in other studies, where dyspnea, increased heart rate and decreased PaO_2_ / FiO_2_ value are evidenced in patients with COVID-19 admitted to the intensive care service ^31^. On the other hand, the median static compliance of this analysis was in the range reported by other studies of 27 to 35 ml/cm H_2_O ^32–34^. However, the high-altitude group showed lower oxygenation values and higher static compliance values, a result similar to the report of the absence of correlation between PaO_2_ / FiO_2_ and static compliance in patients with COVID-19 found by Giacomo et al. ^32^. is a consequence of the fact that the lungs of patients with COVID-19 have vascular characteristics secondary to endothelial damage that differentiates it from the isolated pulmonary involvement of other lung diseases (28).

The treatment basis for moderate-severe ARDS secondary to COVID-19 is based on ventilatory support with low tidal volumes, a prone position, and active management of intravenous fluids ^19,20,35,36^. In this way, it is evident that the strategy The ventilator used for both groups in the present study was protective and up to 79.1% of patients complied with the prone position. Despite these measures, the complications were not different, and the high-altitude group was characterized by higher survival with a median of 39 days, compared to 21 days in the low-altitude group. A difference that remained constant in the subgroup without comorbidities provides one of the first measurements of the contribution of altitude as a factor to be considered in this pathology. It is also important to note that currently the evidence for the use of dexamethasone has been established as a protective factor for mortality ^37^; however, the low-altitude group that received a greater amount of corticosteroids presented higher mortality than the high-altitude group.

In the Cox regression model, 5 factors associated with the risk of death were evidenced. These findings of increased risk do not differ from what is reported in the literature, for its part the APACHE II score has been an effective clinical tool to predict hospital mortality in patients with coronavirus disease 2019 ^38^. Thromboembolic events have also been shown to pose a significant risk of mortality in critically ill patients ^39^. Regarding the SO_2_ / FiO_2_ ratio, it has been widely studied as a prognostic factor ^40^. Besides, as protection, tracheostomy is a procedure that favors the release of ventilatory support in patients with prolonged mechanical ventilation, for which it has been classified as a protective factor against severe complications ^41^. Finally, although altitude has been reported as a possible intervening factor in the clinical outcome of several patients ^7,8^, this is the first study to elucidate more causal information on this relationship.

## Limitations

This study was carried out in two of COVID-19 designed hospitals, part of the social security health system. The private for-profit health system also receives COVID-19 patients, nevertheless, this population was not included in our analysis. To strengthen the findings, there needs to be further study with greater numbers of study sites, greater patient sample sizes and across countries. To better understand the relationship between severity of disease and high-altitude, a range of altitudes needs to be studied, as our study only investigated two altitudes above sea level.

## Conclusion

In this series of cases of critically ill patients with COVID-19 who were admitted to the ICU, there has been a substantial improvement in survival amongst people admitted to the high-altitude critical care unit. High altitude living was associated with improved survival, especially among patients with no comorbidities. COVID-19 patients admitted to the high-altitude ICU unit have improved severity-of-disease classification system scores at 72 hours and reported better respiratory and ventilatory profiles than the low altitude group. Our analysis suggests this improvement is not due to temporal changes in the age, sex or major comorbidity burden of admitted patients.

## Data Availability

Due to the nature of the data and according to local regulation, information concerning patients results cannot be published. Nevertheless, the data that support the findings of this study are available from the corresponding author, Dr. Esteban Ortiz-Prado upon reasonable request at.

## List of Abbreviations

ACE2: Type-2 angiotensin-converting enzyme
ARDS: Acute Respiratory Distress Syndrome
BMI: Body Mass Index
CHF: Congestive Heart Failure
CKD: Chronic Kidney Disease
COPD: Chronic Obstructive Pulmonary Disease
COVID-19: Novel Coronavirus disease 2019
CT: Computed Tomography
CVD: Cerebrovascular Disease
ICU: Intensive care unit
IQR: Interquartile Range
TB: Tuberculosis

## Declarations and acknowledgments

### Ethical Approval

This work was approved by the Hospital IESS-Quito-Sur Internal Review Board (IRB). The request for authorization was submitted in March 1^st^ 2020 and received ethical approval with the following identification number: ID: IESS-HG-SQ-CIE-2020-2656-M.

According to good clinical practices and local regulations, identifiable data from clinical records was only accessed by the medical team that was providing treatment and care to the patients.

### Availability of supporting data

Due to the nature of the data and according to local regulation, information concerning patients’ results cannot be published. Nevertheless, the data that support the findings of this study are available from the corresponding author, Dr. Esteban Ortiz-Prado upon reasonable request at.

### Competing interests

The authors have no conflict of interest to declare

### Funding

This work did not receive a formal grant: however, it received financial support associated with the publication fee from the University of the Americas in Quito, Ecuador.

## Acknowledgements

The authors wish to thank Alberto Sper Sempértegui, Diana C. Guanotoa Muñoz, Carlos P. Pérez Barona, Brayan A. Flores Reyes, Paul X. Garcés Villegas, Eliana M. Morejon Rosero, Josué E. Castro Veintimilla, Rolando J. Chiluisa, Juan C. Jacome Guerrero, Wilson O. Echeverría Mora, Tatiana del Rocío Moreno Paz, Nery M. Cabrera Muñoz, Gerardo D. Zhunio Zhunio, Orlando A. Del Campo Torres, Maria I. Guanga Cadme, Ivonne Z. Peña Escalona, Zulay J. Ochoa Martinez who were very keen in filling the database of each patient recruited in the study.

## Author Disclosure Statement

The authors declare no conflicts of interest.

## Notes

### Competing Interest Statement

The authors have declared no competing interest.

### Author Declarations

This work was approved by the Hospital IESS-Quito-Sur Internal Review Board (IRB). The request for authorization was submitted in March 1st 2020 and received ethical approval with the following identification number: ID: IESS-HG-SQ-CIE-2020-2656-M. According to good clinical practices and local regulations, identifiable data from clinical records was only accessed by the medical team that was providing treatment and care to the patients

